# Machine-Learning Powered Optoacoustic Sensor for Diabetes Progression

**DOI:** 10.1101/2021.03.17.21253779

**Authors:** Pouyan Mohajerani, Juan Aguirre, Murad Omar, Hailong He, Angelos Karlas, Nikolina-Alexia Fasoula, Jessica Lutz, Michael Kallmayer, Hans-Henning Eckstein, Anette-Gabriele Ziegler, Martin Füchtenbusch, Vasilis Ntziachristos

## Abstract

The assessment of diabetes severity relies primarily on a count of clinical complications to empirically characterize disease. Disease staging based on clinical complications also employs a scoring system that may not be optimally suited for analysis of earlier stages of diabetes development or for monitoring smaller increments of disease progress with high precision. We propose a novel sensor, which goes beyond the abilities of current state-of-the-art approaches and introduces a new concept in the assessment of biomedical markers by means of ultra-broadband optoacoustic detection. Being insensitive to photon scattering, the new sensor can resolve optical biomarkers in fine detail and as a function of depth and relates epidermal and dermal morphological and micro-vascular density features to diabetes state. We demonstrate basic sensor characteristics in phantoms and examine the novel sensing concept presented in a pilot study using data from 86 participants (20 healthy and 66 diabetic) at an ultra-wide optoacoustic bandwidth of 120 MHz. Machine learning based on ensemble trees was developed and trained in a supervised fashion and subsequently used to examine the relation of sensor data to disease severity, in particular as it associates to diabetes without complications vs. diabetic neuropathy or atherosclerotic cardiovascular disease. We also investigated the sensor performance in relation to HbA1C values. The proposed method achieved statistically significant detection in all different patient groups. The effect of technical parameters, in particular sensor area size and the time window of optoacoustic signals used in data training were also examined in measurements from phantoms and humans. We discuss how optoacoustic sensors fundamentally solve limitations present in optical sensing and, empowered by machine learning, open a new chapter in non-invasive portable sensing for biomedical applications.

## Introduction

Portable sensors that measure biomarkers indicative of disease are expected to play a critical role in future healthcare [1-8]. Such sensors can be used for frequent measurements of pathophysiological or biochemical parameters on an individualized basis and collect data indicative of disease stage, progression, efficacy of interventions or lifestyle changes. The emerging importance of such biomedical sensors is underscored by growing research on technologies appropriate for biomedical measurements [7] and the corresponding market introduction of wearable or portable devices, which aim to provide disseminated biomedical readings and possibly home-based medical diagnostics or treatment monitoring.

There are different classes of biomedical sensors considered for portable or wearable use. Movement sensors have seen wide commercial deployment and employ a combination of accelerometers and altimeters to measure information such as steps taken, floors climbed, and sleep quality. These sensors are prolific in smartphones and wearable devices, such as Fitbit, the Apple Watch or related products. However, the activity data collected by these devices has only indirect clinical relevance. Wearable devices that track movement may be integrated with electrophysiological sensors that can record electrocardiograms (ECGs), providing data regarding heart rhythm and associated irregularities [9], [10]. Devices such as the Apple Watch’s ECG App or the MC10 BioStamp nPoint have now received FDA clearance for collecting medical grade data with diagnostic potential [11, 12]; a legal recognition that underscores the rapid movement towards proactive healthcare.

Despite successful commercialization, motion and ECG sensors offer limited biomedical information primarily associated with activity, sleep patterns or cardiac function. To extend the possibilities of assessing disease biomarkers in a portable fashion, there has been significant research growth associated with biochemical sensors, i.e. devices that indirectly monitor chemical changes in tissue, typically by sampling biomarkers in extravascular biofluids [13]. Analytes, such as small ions, sugars, steroids and small proteins have been examined in different biofluids, including dermal interstitial fluid (ISF), sweat, saliva and tears. A promising target for non-invasive sensing using wearable biochemical sensors is sweat, possibly stimulated electrically or chemically to achieve consistent flow rates. Compared to other biofluids, concentrations of low molecular weight analytes in ISF appears to correlate better with the corresponding values in blood [14-18]. Nevertheless, concerns about biofluid contamination, low or variable analyte concentration and the precision by which biofluid analyte concentrations relate to those of analytes in blood has so far limited wide applications of biochemical sensing. Wearable biochemical sensors that monitor glucose in ISF are examples that have achieved commercial success. However, it is generally acknowledged that glucose observations in ISF are only indicative and should be confirmed by measurements in blood [7]. An added limitation is that reliable sampling of ISF still requires the insertion of needles and catheters into the skin, making the measurement invasive. Moreover, there is an intrinsic lag-time between analyte concentrations in ISF compared with blood.

A different class of non-invasive sensing is based on optical sensors. Optical measurements are unique in that they sample molecules in a non-invasive manner by recording the modification of a light property by different molecules or tissue structures through the skin surface, i.e. without the need for needles. Several portable or wearable devices now integrate heart rate measurements based on illumination of tissue with green light. Because hemoglobin absorbs green light, variations in the intensity of reflected light relate to blood pulsation and are used to calculate heart rate. Although these measurements are not perfectly accurate, the continuous collection of heart rhythm data allows these devices to possibly alert the user to irregularities. Following the example of the pulse oximeter, some wearable devices also offer measurements of arterial oxygen saturation [19]. Nevertheless, the great challenge of optical sensors relates to photon scattering in tissue, which reduces the accuracy of the physiological data collected and impedes measurements of vascular or morphological features. Therefore, while optical readings offer safe and non-invasive measurements of pathophysiological and possibly biochemical parameters, photon scattering reduces performance and has limited wide application of such sensors in medical measurements.

Aiming for a portable sensor that can go beyond the current state-of-the-art, we propose herein an optoacoustic sensor that retains the non-invasive nature of optical sensors while solving their limitations, in particular their sensitivity to photon scattering. In particular, we developed ultra-wide band (UWB) optoacoustic sensing, that non-invasively detects optical contrast by sensing ultrasound waves generated in response to light absorption by tissue structures [20, 21]. UWB optoacoustics come with two major ground-breaking features over optical sensors. First, optoacoustic sensing has low sensitivity to photon scattering, therefore absorption sources can be accurately detected and quantified with significantly higher precision compared to optical sensing. Second, the use of UWB sensing affords detailed detection of optical contrast as a function of depth. In contrast to optical sensors that offer a single measurement per point, the UWB optoacoustic sensor offers hundreds of measurements of optical contrast along depth. Optoacoustic sensing has been considered in the past as a thermometer [22] or for non-invasive glucose sensing using only superficial measurements in the mid-IR spectral range [23], neither of which utilize the two aforementioned critical features of UWB optoacoustic sensing. Here, we introduce UWB detection as a means of assessing depth-related information, in particular recording features that relate to skin morphology and microvascular density at different depths, as well as vessel size distribution. This type of sensor bridges the gap between optical sensing and advanced imaging methods, offering one-dimensional information along the skin depth. In this way, it uses the skin as a diagnostic window for diabetes progression.

We hypothesized in particular that the one-dimensional measurement of skin optical features using UWB optoacoustic detection would lead to a new class of diabetes progression sensing, useful in diabetes grading. The new sensor differs markedly from the concept of glucose sensing, as it offers a direct systemic measurement of disease progression by quantifying the effect of the disease on the microvascular system; used herein as a novel label-free biomarker of diabetes. In contrast, a glucose sensor detects the level of glucose in blood at the time of the measurement, not the systemic effects of disease. Our hypothesis is based on evidence from several clinical studies, which showed that the status of dermal microvasculature reflects diabetes severity, beginning with early disease development [24, 25]. However, previous studies have performed these observations using histological analysis of biopsy specimens. While histology offers accurate measures of the microvasculature condition, it is not appropriate for frequent and longitudinal measurements due to the invasive nature and laborious analysis required. In contrast, non-invasive sensing of the skin microvascular structure would be highly preferred. A further important advantage of our hypothesis is that there is no other label-free method today that has been shown to be capable of detecting vessel diameters spanning from 5 µm at shallower regions to 100 µm in the deep dermis, i.e. at depths of 1-2 mm or more [26]. For this reason, the use of the skin as a window to diabetes has thus far not been considered.

This development addresses an unmet need in diabetes monitoring. Diabetes is a chronic metabolic disorder that is emerging as a global epidemic [27], and is considered a major threat to human health in the 21^st^ century. Glucose sensors are currently necessary for the more than 400 million people affected by the disease, however glucose monitoring helps with daily disease management and regulating insulin levels in the blood [28], but not with understanding disease status and progression. The ability to monitor diabetes progression in a disseminated and portable manner is critical for efficient management of such large patient pools and could be fundamental to prevention strategies, motivating lifestyle changes or for monitoring the efficacy of therapeutic interventions. Traditional medical tests, such as measuring blood glucose levels after fasting [29] or glycated hemoglobin (HbA1c) analysis can be used for diabetes diagnosis, but these are invasive, laborious and are not well suited to monitor disease progression [30, 31], especially after insulin regulation interventions. We envision herein that frequent sensing of diabetes biomarkers representative of systemic effects and disease progression could improve the assessment of disease severity, now indirectly assessed by major comorbidity events, such as neuropathy, blindness, heart failure, stroke, depression and cognitive dysfunction [24].

The new sensor (Fig. 1a) introduced herein operates with ultrasonic detection at the 10-120 MHZ band (***see Methods***), for precise depth dependent detection of skin vasculature that may reach sub-10 micron layer discrimination, covering the entire epidermal and dermal skin layers. Due to the ultra-wide band utilized, the sensor further collects information from blood (micro-)vessels in the sub-10 micron to 150 micron diameter range, with the smaller vessels yielding contrast at higher ultrasound frequencies, while larger vessels are represented by lower frequencies. The sensor device consisted of a 2-fiber illuminator using 2 ns, 532 nm pulses at a 1 KHz repetition rate. Detection was based on a single ultra-bandwidth lithium niobate LiNBO3 crystal detector (***see Methods***). The sensor was mounted onto a raster scan system (Fig. 1e) for two reasons. First, this arrangement allowed us to investigate the influence of the effective sensing area on the data collected by integrating sensor data from different fields of view. Second, the use of raster scanning allowed us to collect data appropriate for ultra-bandwidth raster-scan optoacoustic mesoscopy (UB-RSOM) [32, 33], so that one-dimensional sensor data could be correlated with skin structures revealed by three-dimensional optoacoustic images, as exemplified in Fig. 1b, obtained from a healthy volunteer. The optoacoustic signal is collected over time, with longer times corresponding to signals coming from deeper in the tissue. Fig. 1c depicts a raw signal obtained by the sensor, corresponding to Fig. 1b. By selecting different time gates (color bars; Fig. 1c) different skin layers can be probed.

**Figure 1.**
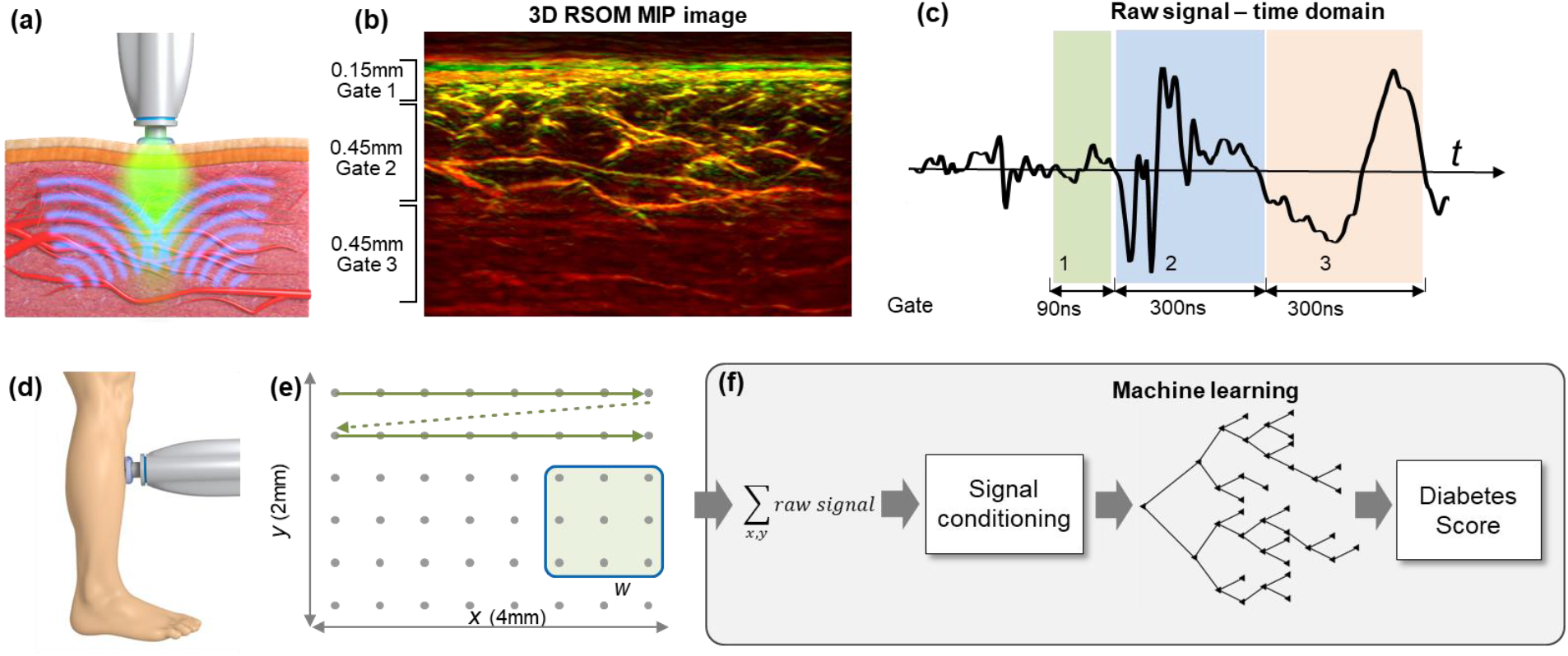
Machine-learning powered optoacoustic sensor. (a) The sensor is placed on the skin and simultaneously emits light into the tissue and collects generated acoustic signals emanating from different depths. (b) A 3D raster-scan optoacoustic mesoscopy (RSOM) reconstructed image, where the small veins dominate the epidermis region (corresponding to gate 1) and large veins are present in the dermis (gate 2 and 3). Gate 2 denotes the first part of the dermis and gate 3 corresponds to deeper regions. (c) A raw signal obtained using the optoacoustic sensor vs. time, where the 3 gates are shown with different colors. (d) A schematic depicting the location on the shin of patients where images are recorded by our in-house sensor system. The acquired time-series data is converted to sensor signals via integration in spatial windows of size *w* × *w*, as depicted in (e). (f) Multiple sensor signals are generated for each patient. The sensor signals from all patients constitute the dataset. The dataset is annotated in terms of diabetes status using patients’ medical history. Machine learning based on bagged ensemble trees is then performed in a supervised fashion to generate predictions about the diabetic status given the 1D optoacoustic sensor signal.

Despite the visual analysis of raw data in Fig. 1c, a particular challenge in analyzing optoacoustic sensing data and testing our hypotheses relates to the rich information content contained in the raw time-resolved optoacoustic sensor signals (Fig. 1c), which offers multiple measurements over depth. This performance is in stark contrast to single point optical sensor data, which only yield a single spatial measurement per point. To process this rich information content of optoacoustic sensing and relate it to the diabetes state, we employed machine learning methodology (Fig. 1f). Machine learning (ML) is well-suited to this task for the following reasons. The raw data from a single-point sensor contains up to a thousand time-points that are not directly interpretable by a human observer (see Suppl. Fig. 1). Machine learning excels at identifying correlations (between data and labels), which are otherwise too complicated for a human observer to discern [34], and generating hypotheses and efficient deterministic tools for data analysis. Conversely, ML is commonly applied to similar streams of data, for example obtained from EEG or ECG [35, 36] [37]. The ML algorithm developed herein consisted of a cascade of unsupervised learning based on principal component analysis (PCA) and supervised learning based on ensemble trees [38] (**see Methods**). The use of PCA points to unsupervised pre-training, aimed mainly at alleviating the adverse effect of the so-called “curse of dimensionality” [39]. The ensemble tree relies on a voting mechanism between learned decision trees, each trained on a resampled training set (with replacement), to achieve lower overfitting and better generalization to unseen data. After training, the ML system assigned a score to each patient (**see Methods**).

To examine our hypotheses and the overall performance of machine-learning powered optoacoustic sensing, we analyzed optoacoustic measurements acquired from a cohort of 86 diabetic patients and healthy volunteers. Measurements were obtained from the front shin area, located 5-10 cm above the medial malleolus area (Fig. 1e). We investigated the UWB sensor in conjunction with diabetic status in 3 scenarios, denoted as Case A, B and C. In **<u>Case A</u>**, the ML algorithm was trained on the 3-class problem of differentiating between 3 groups: healthy (label 0), diabetic without neuropathy (label 1) and diabetic with neuropathy (label 2). In **<u>Case B</u>**, the 3 groups consisted of: healthy (label 0), diabetic without atherosclerotic cardiovascular disease (ASCVD) (label 1) and diabetic with ASCVD (label 2). In **<u>Case C</u>**, the 3 groups consisted of: healthy (label 0), diabetic patients with hemoglobin A1c (HbA1c) values less than 7% (label 1) and diabetic patients with HbA1c values higher than 7.0% (label 2). The number of patients and the age and gender statistics for each subgroup are given in Table 2 in ***Methods***. The differentiation ability of the sensor between the 3 groups of patients in each Case (A, B and C) was analyzed using ANOVA (Analysis of Variance).

**Table 2.** Age, gender and HbA1c for each of the patient groups.

|  | Male | Female | Age, mean | Age, standard deviation | Average HbA1c |
| --- | --- | --- | --- | --- | --- |
| Healthy | 7 | 13 | 30 | 9 | Not measured |
| Diabetic without neuropathy | 11 | 11 | 55 | 19 | 7.1% |
| Diabetic with neuropathy | 32 | 12 | 72 | 9 | 7.1% |
| Diabetic without ASCVD | 26 | 17 | 62 | 16 | 7.2% |
| Diabetic with ASCVD | 17 | 6 | 76 | 7 | 6.9% |
| Diabetic with HbA1c < 7.0 | 21 | 8 | 69 | 15 | 6.3% |
| Diabetic with HbA1c ≥ 7.0 | 22 | 15 | 65 | 16 | 7.8% |

To optimize sensor performance, the ML model was trained for different sensor sizes and time-gates of the data collected (Fig. 1). The sensor size denotes the scan area, or equivalently the skin volume, probed by the sensor. The time-gate relates to the depth layer that is included in the training set, selected by considering signals from a specific time window. The sensor size and time-gate were important to understand the influence of sensor design parameters and also the optimal anatomical site in the skin (skin layer) for obtaining information on diabetes. The sensor size and the time-gate were studied for all three Cases A, B and C.

To corroborate the outcome of the ML analysis on the clinical data, we further conducted two studies, one on phantoms and one using data from human measurements. For the first study, we constructed two phantoms: a simple phantom and a complex phantom. The phantoms were made from 30 μm black surgical sutures (Ethicon, Inc. USA). The sutures were submerged in a water bath mixed with a 1 % solution of intralipid 20% (Sigma-Aldrich, USA) to obtain a slightly scattering medium. The simple phantom contained two sutures to simulate low vascularization.

The complex phantom contained five futures to simulate denser microvasculature. The phantoms were scanned with an identical procedure to the one used for the human measurements. For the second study, we contrasted sensor data against corresponding optoacoustic images of skin microvasculature in order to better understand the basis of the contrast in the data.

## Results

The initial investigation into sensor performance was directed toward identifying whether raw sensor data captured information that was so far available only from optoacoustic images. Phantom measurements were rendered as images (Fig. 2a,b) and as raw sensor data (Fig. 2c). Visual inspection of raw sensor data revealed that signals from the complex phantom showed large intensity variations as a function of time (depth) at depths corresponding to the presence of the sutures, whereby signals from the simpler phantom varied less due to the smaller number of sutures. The finding shows that variation in contrast clearly seen in the images is also reflected in raw sensor data. To investigate the sensor ability to differentiate between the simple and complex phantoms as a function of sensor size we performed dimensionality reduction based on t-SNE [40], applied on the time-series signals (Fig. 2d-h). The separation between point clouds, corresponding to the simple phantom (red) and the complex phantom (gray), increases with sensor size. Quantification of the distance between the two cloud groups (Supp. Fig. 2) supports this visual observation and confirms that the differentiation ability increases with larger sensor area.

**Figure 2.**
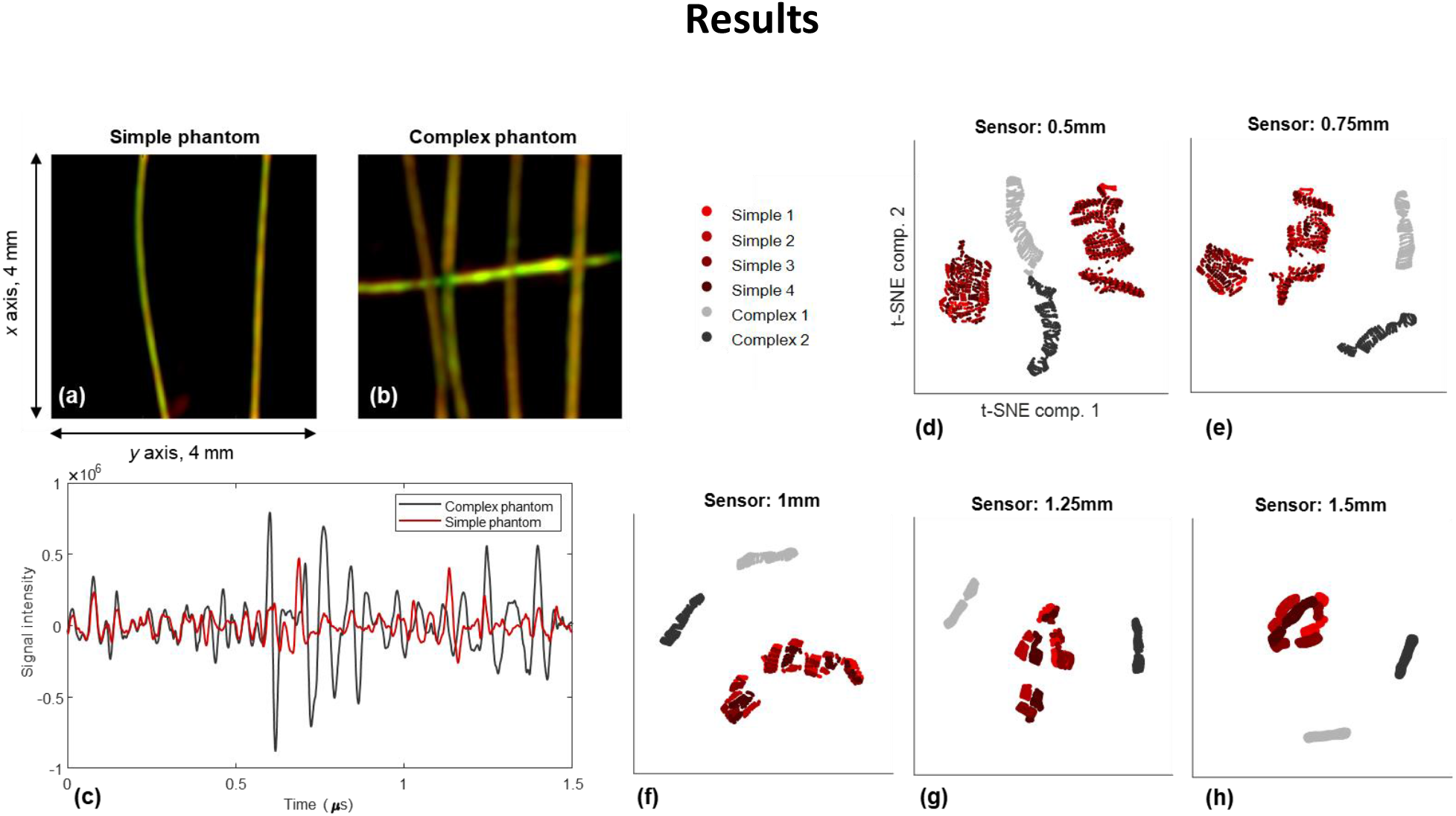
Feasibility demonstration using optoacoustic phantoms. Panels (a) and (b) show top-view RSOM images from the simple phantom with two sutures and the complex phantom with five sutures, respectively. Sensor signals from the two phantoms are shown in (c). (d-h) show, for different sensor sizes, signals after dimensionality reduction in the t-SNE domain. Red points correspond to four repeated measurements from the simple phantom and the gray points correspond to two repeated measurements from the complex phantom.

To examine whether the findings from phantoms translate to measurements from humans, we further analyzed sensor signals from healthy and diabetic skin (Fig. 3). The RSOM image from the healthy skin (Fig. 3a) visibly shows denser vasculature than the diabetic skin (Fig. 3b), particularly in the dermal region. Observation of sensor raw data (Fig. 3c) shows that, similar to the phantom signals, the healthy skin exhibits stronger intensity variations than the diabetic skin. These variations are more prominent deeper in the skin, consistent with the appearance of vascular density in the images. To examine whether this observation is consistent with the hypothesis of optoacoustic contrast due to declining vascular density in the diabetic skin, we further analyzed the signals in the frequency domain (Fig. 3d,e). Frequency analysis of the raw time signals describes the sensor’s ability to measure the distribution of blood vessel size, since smaller vessels emit higher ultrasound (optoacoustic) frequencies compared to larger vessels. Intensity normalized frequency profiles, corresponding to the signals in Fig. 3c, exhibited different spectral characteristics in the dermis and epidermis between the healthy (black curves) and the diabetic skin (red curves). In both skin layers, healthy skin exhibits a spectrum containing higher frequencies than the diabetic skin. This observation can be attributed to loss of finer vasculature in the diabetic, leading to a corresponding reduction of the high-frequency content in the sensor measurement.

**Figure 3.**
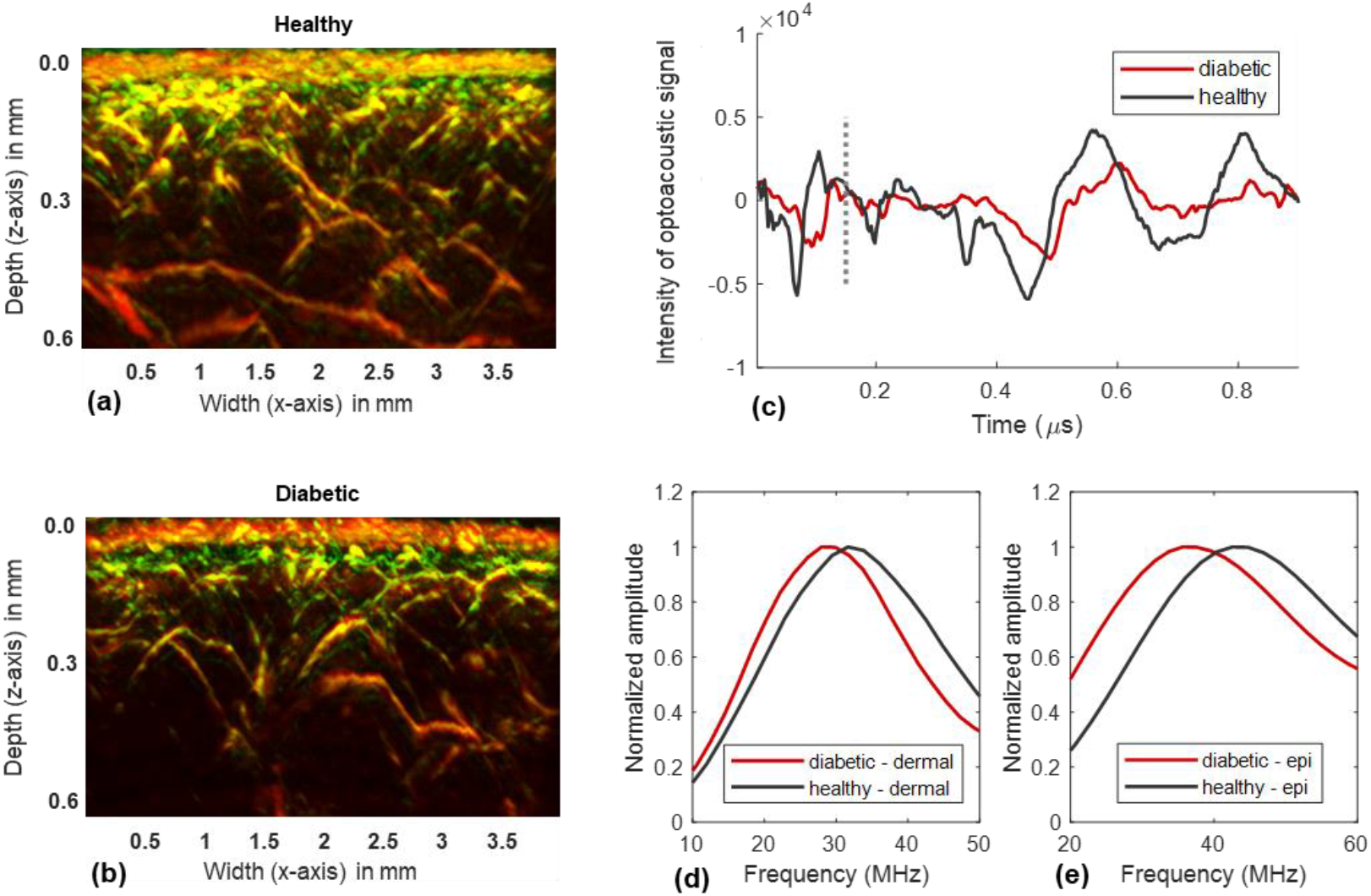
Sensor signals and images from a healthy volunteer a diabetic patient. Panels (a) and (b) show maximum intensity projection (MIP) images from RSOM reconstructions for a healthy volunteer and a diabetic patient, respectively. Raw time-series sensor signals from the two participants are shown in (c), where the healthy signal (black) manifests a larger dynamic range than the diabetic signal (red). Panels (d) and (e) show signals in the frequency domain of the dermis and epidermis, respectively.

While the results presented in Fig. 3 exemplify the differences between signals from the healthy and diabetic skin, such differences become subtler and difficult to distinguish as we observed the progression of disease, i.e. when we examine patients with different diabetes stage (see Supp. Fig. 1). A key goal of the study herein was not to provide evidence of microvascular differences between the healthy vs. the diabetic skin, but to study whether optoacoustic contrast collected by a sensor relates to disease state. For this reason, subsequent analysis of the data obtained from humans was performed using machine learning.

Results from the human measurements in the cohort are presented for four different sensor sizes of 0.3 mm, 0.6 mm, 1.2 mm and 1.8 mm, which represent realistic scenarios for a portable sensor design. Additionally, three time gates were considered, as shown in Fig. 1b,c. The first gate starts at the skin surface and extends for 0.075 µs or 0.12 mm. For reference, the epidermis has an average thickness of 0.1 mm while the dermis layer has a thickness of up to 2 mm [26]. Gates 2 and 3 follow after Gate 1 and are each 0.45 mm (0.1 µs) deep and cover the upper and lower parts of the dermis. Therefore, Gate 1 corresponds to the epidermis layer, Gate 2 corresponds to the connecting vessels in the upper part of dermis and Gate 3 corresponds to lower vascular plexus in the deeper portion of the dermis.

Fig. 4 shows the performance of the proposed system for the three cases that were examined. The top, middle and bottom rows in Fig. 4 depict the predicted scores for Case A, Case B and Case C, respectively, in the form of bar plots. The black dots show the average scores for the respective patient groups and the length of the vertical bars correspond to double the standard deviation of the predicted scores. For each Case, the bar plots for the three classes are depicted with blue, green and red colors. The left panels (Fig. 4a,c,e) show the scores versus the four sensor sizes, whereas the right panels (Fig. 4b,d,f) depict the scores when different time-gates are used for the largest sensor size (1.8 mm).

**Figure 4.**
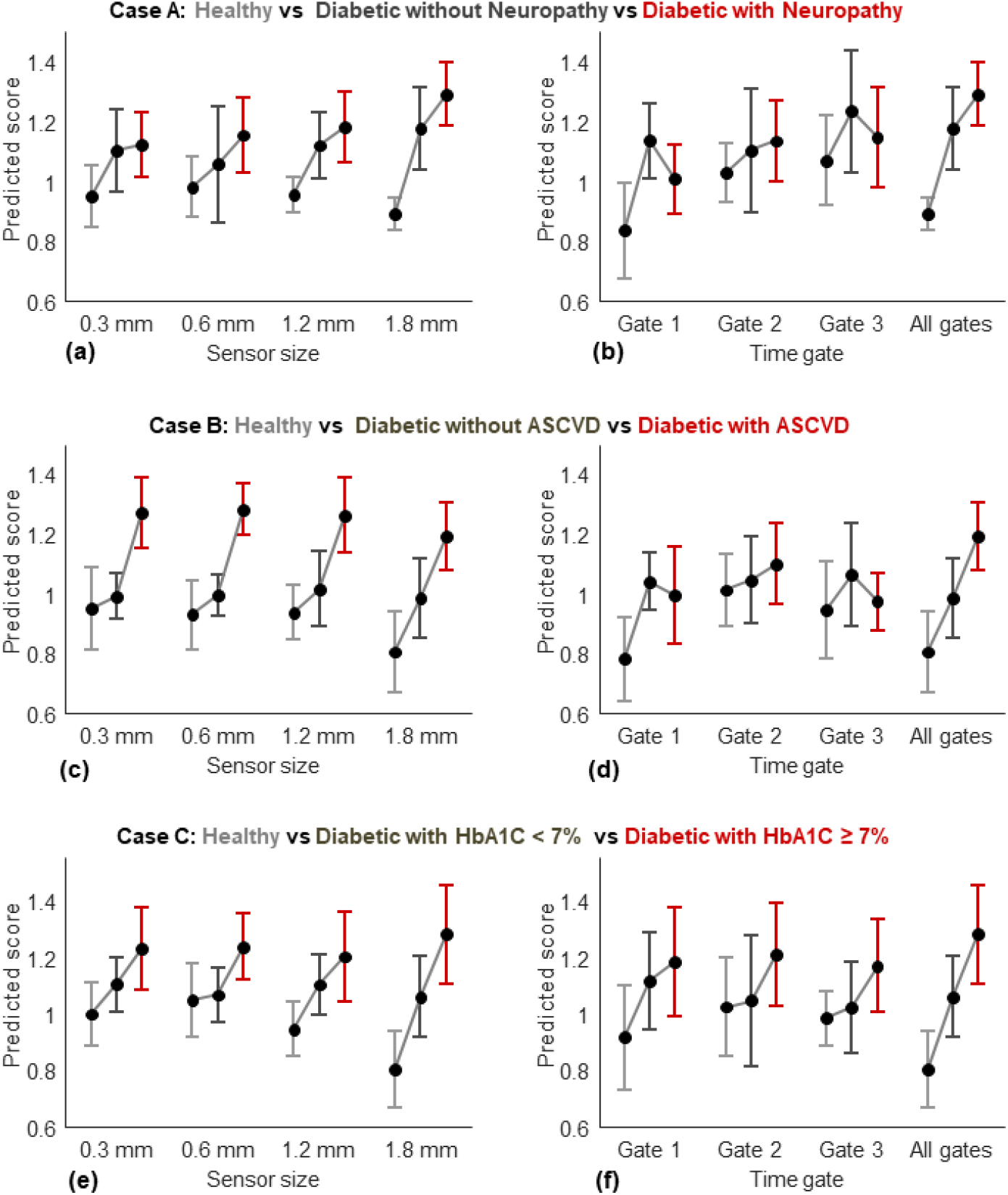
(a) Bar plots show the predicted scores vs. sensor size (w in Fig. 1f) for different groups of patients (as denoted in the title by the colors blue, green and red) for Case A. The bottom and top parts of the bar plots show the corresponding standard deviation (therefore, the total vertical length is equal to double the standard deviation). An increasing trend (shown by the black line connecting the black dots) in signal vs. disease status is desirable. (b) The predicted scores vs. time gate, where gate 2 is seen to have the most amount of predictive information. (c, d) The results for Case B vs. sensor size and time gate, respectively. Similar to Case A, the second time gate contains the most information. (e, f) The corresponding results for Case C, where the 2nd time-gate again has the most information. The differentiation improves in Case C as the sensor size is increased. The number of patients in each subgroup for each case, is presented in Table 2 in Methods.

For Case A (differentiation of healthy vs. diabetic without neuropathy vs. differentiation with neuropathy), the mean scores (black dots in Fig. 4a) increase in relation to disease presence for all four sensor sizes, but exhibit the best differentiation for the largest area sensor. This result can be explained by considering that a larger sensor area effectively collects more signal from the skin and yields a better signal-to-noise ratio and wider sampling of skin features. The performance was quantified via ANOVA analysis in the form of p-values. The ANOVA p-values for all three cases and all sensor sizes (when using the entire time gate) are tabulated in Table 1. The p-value of 0.004 for the largest sensor in Case A substantiates reasonable differentiation.

**Table 1.** Group differentiation in term of p-value for the 3 Cases A, B and C vs. sensor size (when using all time gates). The largest sensor has the lowest average p-value across all three cases. The p-values were obtained using the one-way ANOVA test.

|  | 0.3 mm | 0.6 mm | 1.2 mm | 1.8 mm |
| --- | --- | --- | --- | --- |
| Case A | 0.159 | 0.231 | 0.121 | 0.004 |
| Case B | 0.002 | 0.001 | 0.006 | 0.011 |
| Case C | 0.062 | 0.087 | 0.061 | 0.002 |

Inspecting now the sensor performance as a function of the gate employed, we see that the best differentiation was achieved when the entire signal was utilized, i.e. all gates. This finding was derived from a sensor with a size of 1.8 mm. When one utilizes independent gates, it is found that only Gate 2 preserves the differentiation between patient groups when used alone. These findings suggest that, as expected, the larger differentiation ability of the sensor comes from Gate 2, i.e. the gate that corresponds to the upper vascular plexus of the dermis. This layer presents the most prominent changes as a function of diabetes severity, i.e. loss of vascularization associated with diabetic progression. Nevertheless, even if the two time gates 1 and 3 do not appear to achieve differentiation on their own, the best differentiation is achieved when all of the time signal is employed as input to the ML algorithm. This result therefore indicates that in addition to dermal vasculature, there are other features in the skin, possibly associated with the overall morphology and deeper vessels, which also contain information on diabetes progression.

Analysis of Case B (Fig. 4c,d) shows a similar behavior for the UWB optoacoustic sensor data. Differentiation is best for the larger sensor area and when all time gates are employed. The p-values for differentiating between healthy, diabetic without ASCVD and diabetic with ASCVD for the different sensor areas are also tabulated in Table 1. In this case, all four sensor sizes achieve p-values under 0.011.

Analysis of Case C (healthy, diabetic with HbA1c < 7% and diabetic with HbA1c ≥ 7%) also follows the same trend as in Cases A and B. Case C examines the relation between microvascular changes and HbA1c, and is indicative of two seemingly independent parameters associated with damage to the vascular system vs. recent history of hemoglobin glycation. The results follow similar trends to those of Cases B and C, i.e. for all sensors the mean values correlate with disease state and gate 2 achieves the best performance. As seen from the p-values in Table 1, the largest sensor achieves the lowest p-value. Further complementary performance analysis results are given in the ***Supplementary material***.

## Discussion

Biomedical optical sensors offer an ideal measurement system by combining three major strengths. First, they offer direct measurements of tissue pathophysiology, since they are based on photons that interact directly with molecules and structures of interest in the tissue examined. This is in contrast, for example, to biochemical sensors, which are invasive and only indirectly assess parameters of tissue by measurements in accessible biofluids. Second, they are non-invasive and use safe energy, therefore they are appropriate for frequent or continuous measurements. Finally, they can be implemented in portable formats and can measure at multiple sites in the body. However, a major caveat of optical sensors so far has been the difficulty of providing accurate measurements, due to the complications of analyses performed with diffusive photons generated by the strong photon scattering in tissue. Optoacoustic sensors address this critical challenge by utilizing ultrasound detection of optical contrast and minimizing the sensitivity of the measurement to scattering. Moreover, the use of UWB detection allows precise localization of signal with depth, offering information never before available to optical sensors. We illustrate in this paper how UWB optoacoustic sensors can surpass simple lifestyle measurements afforded by current optical devices and yield features that can correlate directly with disease.

In particular, we introduced a sensor technology that is based on broadband optoacoustic detection (>100 Mhz bandwidth). Frequency responses due to optoacoustic sensing from the skin span several tens of MHz (See Fig. 3) and a broadband sensor ensures that sufficient information is collected. Signals of broad ultrasonic spectrum captured detailed depth-dependent information on skin morphology and microvasculature, i.e. parameters that have never been previously assessed by a sensor in a non-invasive manner. Consequently, the novel sensor technology enabled the assessment of skin features known to correlate with diabetes progression. By processing the data with machine learning, we have shown that the sensor identifies diabetic patients and also yields a good correlation with the HbA1c levels, as well as disease status for neuropathy and ASCVD. Generally, use of the entire time signal yielded the most prominent differentiation between the examined cases. However, most useful information was found to reside within the depths associated with gate 2, correlating to the upper vascular plexus of the dermis. This finding further supports the use of a depth-discriminating sensor over optical or optoacoustic sensors that either detect only superficial signals or offer a low resolution recording of depth-related features.

There are several implications relating to the introduction of the new sensor. Diabetes progression and severity is only assessed today in an infrequent manner and usually follows the development of late stage clinical symptoms, which prompt a diagnostic test. The availability of a sensor that could monitor changes associated with diabetes progression alters the concept of a diagnostic test. Rather than the disease being diagnosed at a single point in time, frequent measurements enable a continuum of signs to be monitored, which indicate the progression of the disease. In this role, the sensor could be implemented in homebased monitoring, in analogy to today’s ability to monitor lifestyle conditions using motion and electrical sensors. Of particular importance, is that persons monitored may serve as their own reference baselines, i.e. the sensor may prove to be more accurate when relative changes are monitored as a function of time on the same individual.

Assessment of skin microvascular morphology at depths of up to several millimeters at single vessel resolution is desirable, but not attainable by current methods. Pure optical methods like dermoscopy, reflectance confocal microscopy, two-photon microscopy, laser Doppler imaging and laser speckle contrast imaging are fundamentally limited by photon scattering and are incapable of high resolution imaging beyond a few hundred micrometers [41-44]. Optical coherence tomography uses the so-called “coherent gating technique” to overcome the scattering curse, providing morphological images of the skin at resolutions similar to those of RSOM. However, when imaging vascular networks using flow-sensitive techniques in the visible range, penetration is limited to depths of ∼400 μm. Moreover, inherent image artefacts in the axial direction compromise the ability of optical coherence tomography to obtain cross-sectional images [45-48]. Therefore, optoacoustic methods could offer better microvasculature imaging performance compared to OCT [50, 51]. Pure ultrasound imaging techniques can penetrate several millimeters into skin tissue. However, the contrast mechanism, which is based on sound reflection, does not resolve vessels less than 100 micrometer in diameter unless extrinsic contrast is used, imposing limitations for routine longitudinal applications in humans [49]. In all cases, sensor measurements are more cost- and usage-efficient compared to imaging, especially for enabling high device dissemination [24, 52] [53].

Overall, the ability of the new sensor to offer readings that relate to metabolic disease, beyond the current state-of-the-art sensors, offers the possibility of incorporation into both homecare environments and point-of-care settings. Assessment of diabetic progression after diagnosis is currently based on infrequent testing and assessment of crude clinical features, such as comorbidities (neuropathy, nephropathy etc.). Overall, the number and the severity of complications such as retinopathy, nephropathy, cardiovascular disease, stroke, peripheral arterial disease or neuropathy increase the mortality and hospitalization risk. The adapted Diabetes Complications Severity Index (DCSI) was designed to assess diabetes via its complications, their severity and laboratory data and to provide an estimation of severity [54, 55]. The Diabetes Severity Score (DSS) offers an alternative metric based on parameters, such as the age, the BMI, the duration of diabetes, the presence of microvascular and macrovascular complications, the need for insulin treatment and the levels of stimulated C-peptide in blood [56]. The new sensor provides a measurement of the systemic effect of diabetes using a quantified score, which can be performed in a frequent manner. This data could be considered as a quantitative measurement to complement the information included in the DCSI or DSS, or as a measurement to provide finer assessment of progression within the indices employed, which cannot be captured by the development of clinical symptoms that occur at more infrequent time points.

The pilot study herein outlined the ability of the sensor to capture diabetic progression with patients of varying severity, having different diabetes complications, including neuropathy, ASCVD and different levels of HbA1C values. A particularly interesting next step would be to deploy the sensor in a prospective longitudinal study to examine whether each person monitored may serve as their own reference, i.e. evaluate the sensitivity of the sensor to identify disease progression when relative changes are recorded as a function of time on the same individual. Moreover, we note that while a healthy group was included for comparison, this group was not age-matched and it is only shown here to indicate the sensor baseline. A follow-up study will include age-matched healthy volunteers as well in order to better assess the sensitivity of the sensor to disease related vs. age related micro-vascular changes. Another important future step would be to examine the sensor performance in relation to the number of wavelengths employed for tissue illumination, a relatively straightforward hardware upgrade. Another future goal is the inclusion of other medical information in the ML algorithm, for example demographic information (such as age and sex), anthropometric measurements (such as weight, BMI and waist-hip-ratio), as well as clinical or laboratory measurements. Furthermore, in this work we decided to measure the skin of the lower extremity, relying on the large number of optical sensors studies indicating skin microvascular impairment in this region. Nevertheless, optimization of the measurement location is a topic to be further investigated in the future studies.

It should be noted that although several components of the optoacoustic system might lend themselves to miniaturization, we believe the proposed sensor may be indeed designed to be used in a wearable manner. The ultrasound transducer has a very small form factor and can be easily attached to the skin in a wearable manner. The rest of the components (laser, optics, electronics, power supplies) are placed away from the sensor and only need an optical fiber and a coaxial cable for connection with the transducer. Moreover, such components can exhibit a small form factor. Therefore, while making the system as wireless wearable item can be challenging, it can be assuredly designed to be wearable for studies involving any kind of activity. More in particular, the ultrasound transducer used in the paper is manufactured with a length of 20 mm and height of 7 mm. Furthermore, we have demonstrated that overdriven CW diode laser can be used to perform ultrawide-band optoacoustic imaging of human skin microvasculature [57]. Such lasers, together with the driving electronics are available in a very small form factor. Regarding acquisition electronics, there exist small enough analogue-to-digital convertors with the appropriate sampling rate that can be connected to mobile devices.

In conclusion, the pilot study herein introduced a new sensor concept and demonstrated its ability to use skin and microvascular features for identifying systemic effects of diabetes. This tool is geared toward disseminated use in monitoring diabetes progression and efficacy of possible interventions in a frequent and portable manner. We anticipate that optoacoustic technology will lead to a new paradigm in biomedical portable sensing, finding broad applications for metabolic and cardiovascular diseases.

## Methods

### Sensor data generation and labelling

Optoacoustic signals are acquired on a 533×135 grid in the *x*-*y* plane, with scan step sizes of Δ*x* = 7.5 µm and Δ*y* = 15 µm, resulting in a total scan area of 4 mm × 2 mm. The time-series data acquired at every location contains at least 1000 data points (the actual number varied slightly during the course of the project; accordingly, all time-series measurements were clipped to the first 1000 points). For each measurement, we found the approximate beginning position of the skin automatically using the raw data. The signal before the skin position was set to zero (as it contains noise and reflection) and the first 200 zero values were also discarded in all cases. At a sampling frequency of 1 GHz and a nominal sound speed of 1540 m/s in biological tissue, the remaining 800 data points correspond to 0.81 µs or, equivalently, 1.23 mm.

This work proposes the application of optoacoustic sensors for the diagnosis of diabetes. The sensor signals were simulated from the time-series scan measurements by integration of the time-series measurements (consisting of 800 points as explained above) with a *w* × *w* spatial windows as shown in Fig. 1e. Four different sensor sizes with *w* = 0.3 mm, 0.6 mm, 1.2 mm and 1.8 mm. For each *w*, the window was shifted horizontally and vertically in 0.2 steps and for each position of the sensor window a sensor signal was generated. Given the total scan size of 2 mm × 4 mm, and a window shift of 0.2 mm, the largest sensor size was set to 1.8 mm. With these configurations, and given two imaging locations per person, a total of 420, 304, 160 and 52 sensor signals per person were obtained for sensor sizes 0.3 mm, 0.6 mm, 1.2 mm and 1.8 mm, respectively.

Each sensor signal was labelled using the medical history of the corresponding patient. Accordingly, three classification problems, denoted as Cases A, B and C, were formulated and studied in this work (see Table 2 in Methods for number of patients in each group).

**Case A** – [Label 0: Healthy] vs [Label 1: Diabetic without Neuropathy] vs [Label 2: Diabetic with Neuropathy]

In this case, the first group consists of healthy volunteers. The second and third groups consist of diabetic patients with and without diabetic neuropathy, respectively.

**Case B** – [Label 0: Healthy] vs [Label 1: Diabetic without ASCVD] vs [Label 2: Diabetic with ASCVD]

Similar to Case A, this case considers presence or absence of Atherosclerotic Cardiovascular Disease (ASCVD).

**Case C** – [Label 0: Healthy] vs [Label 1: Diabetic with HbA1C < 7.0%] vs. [Label 2: Diabetic with HbA1C ≥ 7.0%]

In this case, we created a three-class classification problem by dividing the patients into three groups based on their hemoglobin A1c (HbA1c) values. HbA1c denotes the percentage of hemoglobin that is glycated and is the most important measure of chronic glycaemia [58]. A healthy person with no history of diabetes or impaired glucose tolerance has an Hbac1 value of less than 5.7% [59]. Abnormally high values of HbA1c in diabetes signify poor control of blood glucose levels.

### The machine learning workflow

We first describe the notation used herein to describe the data. Let 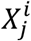 be a 1 × *D* vector representing *j*^*th*^ the integrated sensor signal obtained from subject number *i*. For every subject, *j* ranges from 1 to *J*, where *J* is determined by the scan size and the integration window size *w* (as described above). *D* is the sensor signal length with the value of 810, corresponding to 0.81 µs or, equivalently, 1.24 mm. Furthermore, let *Y*_*i*_ be a categorical variable denoting the diagnostic label for subject *i*. The labels take values 0, 1 or 2, as defined above for the 3 cases.

The goal is to design a machine learning algorithm that takes 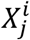 as the input and generates a prediction of the true class *Y*_*i*_. The proposed machine learning pipeline is depicted in Fig. 5. Several machine learning algorithms were examined, with and without preprocessing of sensor data and using deep or classical classifiers. It was observed that bootstrap-aggregated (bagged) decision trees [38] preceded by principal component analysis (PCA) [39] yield the best results.

**Figure 5.**
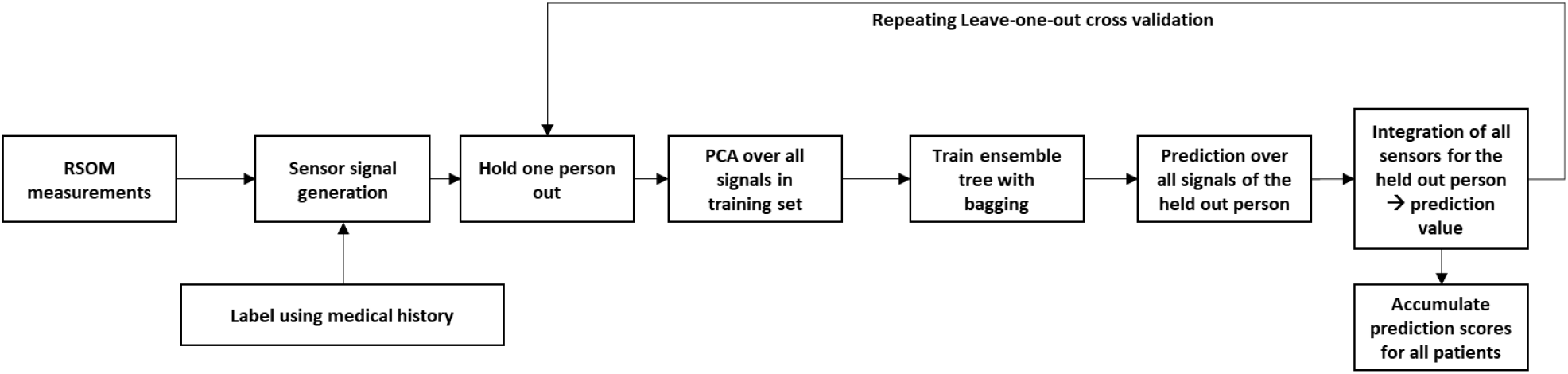
The machine learning pipeline consisting of an unsupervised component (PCA) and a supervised component (bagged ensemble trees). The dataset is obtained from the scan measurements and the labels are extracted from the medical history. The calculation of the PCA matrix as well as the training of the tree ensemble is performed on the training set only. Leave-one-out cross validation (LOOCV) is performed by repeating the training and evaluation process over all the patients and volunteers. For every person in the test set, the predictions are calculated for the trained tree ensemble on all the sensor signals. The average value of these predictions, called the predicted score, gives the final prediction value for the given person.

The combination of PCA and bagged ensemble trees is a form of semi-supervised learning approach and where the PCA is used for dimensionality reduction in unsupervised learning mode and the ensemble tree for the subsequent classification (supervised learning). Such combinations are indeed very common and has been used in many applications [60-62]. PCA removes redundancies in the data and hence improves the performance of the subsequent classification module by reducing the adverse effect of the so-called Curse of Dimensionality. While deep learning generally aims to reduce or remove the need to preprocessing by directly learning useful representations, its application often necessitates large datasets. Transfer learning also requires existence of large datasets of similar nature. Such conditioned are not however fulfilled for our data. Indeed, using PCA to improve performance of deep learning is also common practice Even today PCA is used along with deep learning to improve performance [63-65].

The decision tree model was implemented using MATLAB’s TreeBagger function. Our particular approach is as follows. The goal of the ML system is to generate a prediction score for each patient. The prediction score is a real-valued number between 0 and 2, which should be ideally equal to the label associated with the given patient. We generate this score for the subject *i*, in the following way.

First, for the training set *T*_*i*_ (consisting of the pooled data of all subjects except subject *i*), all of the sensor signals are stacked in a design matrix. PCA is performed on this matrix and the first 15 components are retained. The PCA vectors were normalized to their respective first components. The PCA matrix found from the training set *T*_*i*_ is denoted by *U*_*i*_. The matrix *U*_*i*_ is applied to all samples in the training and evaluation sets (where the evaluation set consists of all the sensor signals from subject *i*). Note that by calculating the PCA matrix from the training set only, we avoid any risk of overfitting.

The bagged ensemble tree is then trained on the vectors in *T*_*i*_. The ensemble consists of 100 trees, with a minimum leaf size set of the number of samples in *T*_*i*_ divided by 30. A very small minimum leaf size leads to overfitting, while a very larger one might lead to bias. No data augmentation was used during the training or the evaluation phase.

Eventually, to obtain the prediction score for patient *i*, the same PCA matrix *U*_*i*_ (obtained from the training set) is applied to the sensor signals of patient *i*, that is 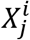. The output rows of this matrix multiplication are then normalized by the first component and the result is fed to the ensemble tree for prediction. The predicted score is then generated as the average of the predictions over all 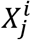 s. Note that averaging model predictions for different inputs (here, sensor signals) extracted from a given test sample (here, the patient) is common practice. An example of such approach is test-time augmentation (TTA), where a single model is used to predict labels over different augmented versions of the same image to obtain a final prediction.

The algorithm’s performance was evaluated for each time gate by setting the signal outside the time gate to 0. The trees were then trained using these truncated signals. The matrix *U*_*i*_ was also used for evaluation of the individual time gates.

Evaluation of the proposed algorithm was performed using Leave-one-out cross-validation (LOOCV), as shown in Fig. 5. That is, from *N* patients, *N*-1 is used for training and 1 is used for testing. This way, the algorithm is tested based on patient data that it has not seen during training. This way, the risk of overfitting is reduced. Using LOOCV for cross-validation and testing of ML algorithms is particularly relevant when the dataset is small, as is the case in our work.

### Clinical imaging

A total of 21 healthy volunteers and 71 patients with a history of diabetes were recruited in this study. Patients were recruited in this single-center study, at the Diabetes Center Munich. All patients and healthy volunteers signed an informed consent approved by the local ethics committees of Helmholtz Zentrum München, as well as the Technical University of Munich.

Subjects were placed in a room with normal temperature (23°C) and measured in supine position. Each patient was scanned with the RSOM system [32, 33], as shown in Fig. 1, over two positions on the distal pretibial region of both lower extremities. These positions were selected as representative of the skin microvasculature, as the skin of the lower extremities is often influenced in the natural course of diabetes. Furthermore, this imaging location supported patient comfort and reduction of motion artefacts during imaging. Each measurement lasted around 70-80 seconds. The measurements were performed during a period of one year from June 2017 to June 2018.

The data from 6 patients were excluded from this study due to bad quality of the reconstructions, arising from excessive motion during imaging. The amount of motion was quantified using an in-house algorithm which measures the curvature of the reconstructed skin surface as an indicator of motion [66]. Consequently, the data from a total of 20 volunteers (7 males, 13 females, age 30 ± 9 years) and 66 patients (43 males, 23 females, age 66 ± 15 years) were pooled in the sensor signal dataset. Further statistics about all patient subgroups are presented below in Table 2.

HbA1c of the patients with diabetes was ∼7.07% in average (average HbA1c values for different patient groups can be found in Table 2). Around one third of the patients included qualified as ASCVD patients. Such patients either had coronary artery disease, carotid artery disease or peripheral arterial disease. The peripheral arterial disease patients showed clinically relevant stenosis detected by Doppler ultrasound measurements. All of these patients had undergone a revascularization procedure. The rest of the ASCVD patients (coronary artery disease and carotid artery disease) had had a cardiovascular event.

Two third of the patients had neuropathy, where the presence of neuropathy was assessed based on neuropathy symptoms score (NSS) and neuropathy disability score (NDS). These scores were chosen due to the ease of assessment for all patients.

### Performance evaluation

For every subject a predicted score *p*_*i*_ was obtained as described above. The one-way analysis of variance (ANOVA) test was used to quantify the differentiation between the three groups. ANOVA is a hypothesis test, were the null hypothesis asserts that the data (here, scores) come from the same population [67]. ANOVA calculates one p-value for all 3 groups. A smaller p-value indicates a higher confidence that the scores are statistically different between the 3 groups.

## Data Availability

Data is available from the authors upon request.

## Acknowledgements

This project has received funding from the European Research Council (ERC) under the European Union’s Horizon 2020 research and innovation programme under grant agreement No 694968 (PREMSOT) and from the Deutsche Forschungsgemeinschaft (DFG), Germany [Gottfried Wilhelm Leibniz Prize 2013; NT 3/10-1]. Furthermore, Ludwig Englert at IBMI is acknowledged for his assistance with obtaining measurements.

## Supplementary Material

**Supplemental Figure 1.**
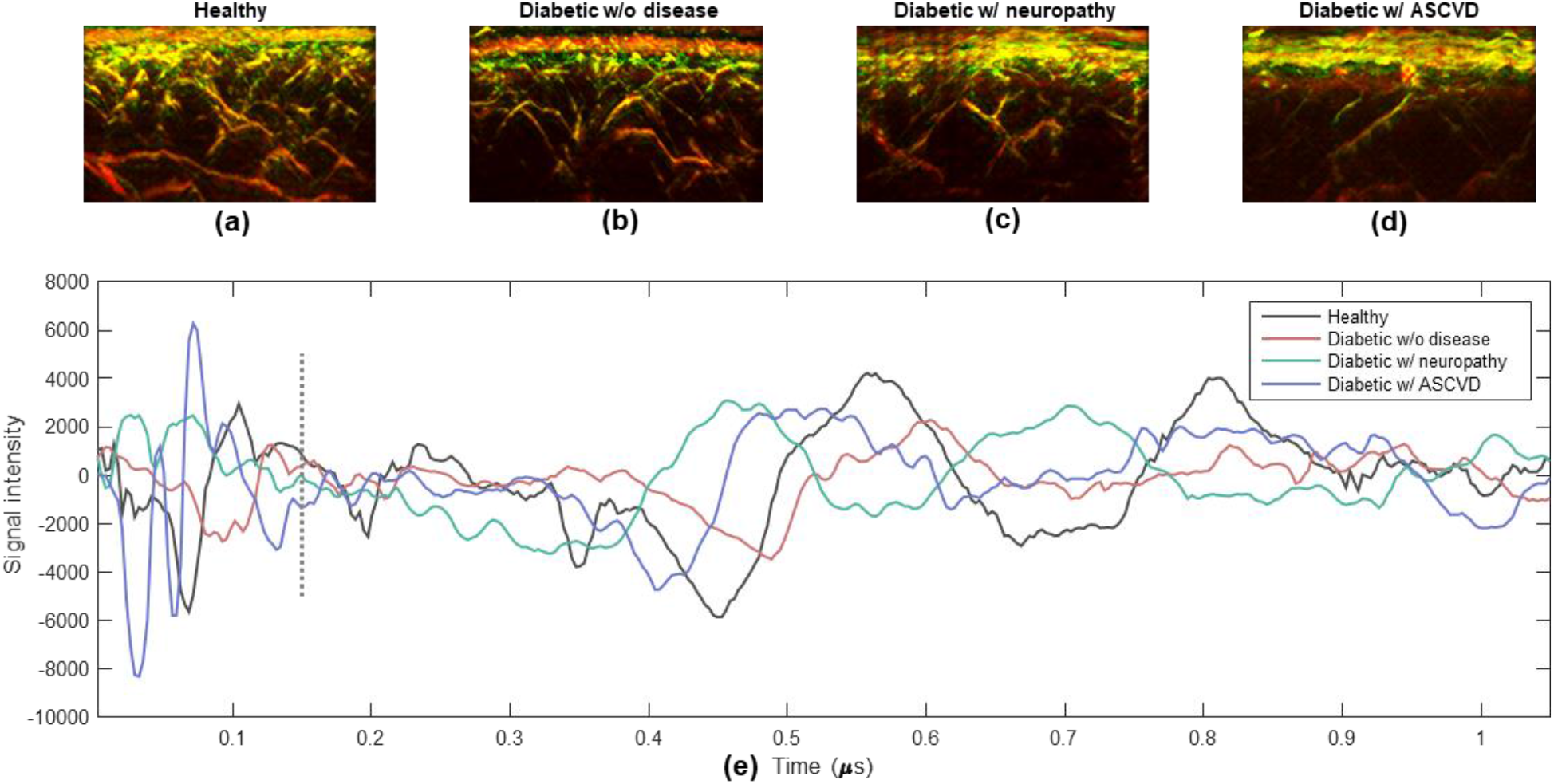
Sensor signal vs. disease progression. Panels (a-d) depict MIP RSOM images for, respectively, a healthy volunteer, a diabetic patient without disease, a diabetic patient with neuropathy and a diabetic patient with ASCVD. Panel (e) shows time-series sensor signals for the four participants.

In Fig. 3 we showed sample signals from healthy and diabetic skin we contrasted them against the RSOM images. In Supp. Fig. 1 we show signals and corresponding RSOM images versus disease progression. The vasculature appears to become sparser as the disease progresses in the top row. However, the differentiation between signals from the four disease groups, shown in Supp. Fig. 1e, does not lend itself to methods based on simple metrics, such as intensity. Therefore, data-driven methods based on machine learning are required to perform the task of signal differentiation and disease staging. As disease progresses, differentiation of signals becomes more complicated, prompting the need for machine-learning algorithms to perform the task.

**Supplemental Figure 2.**
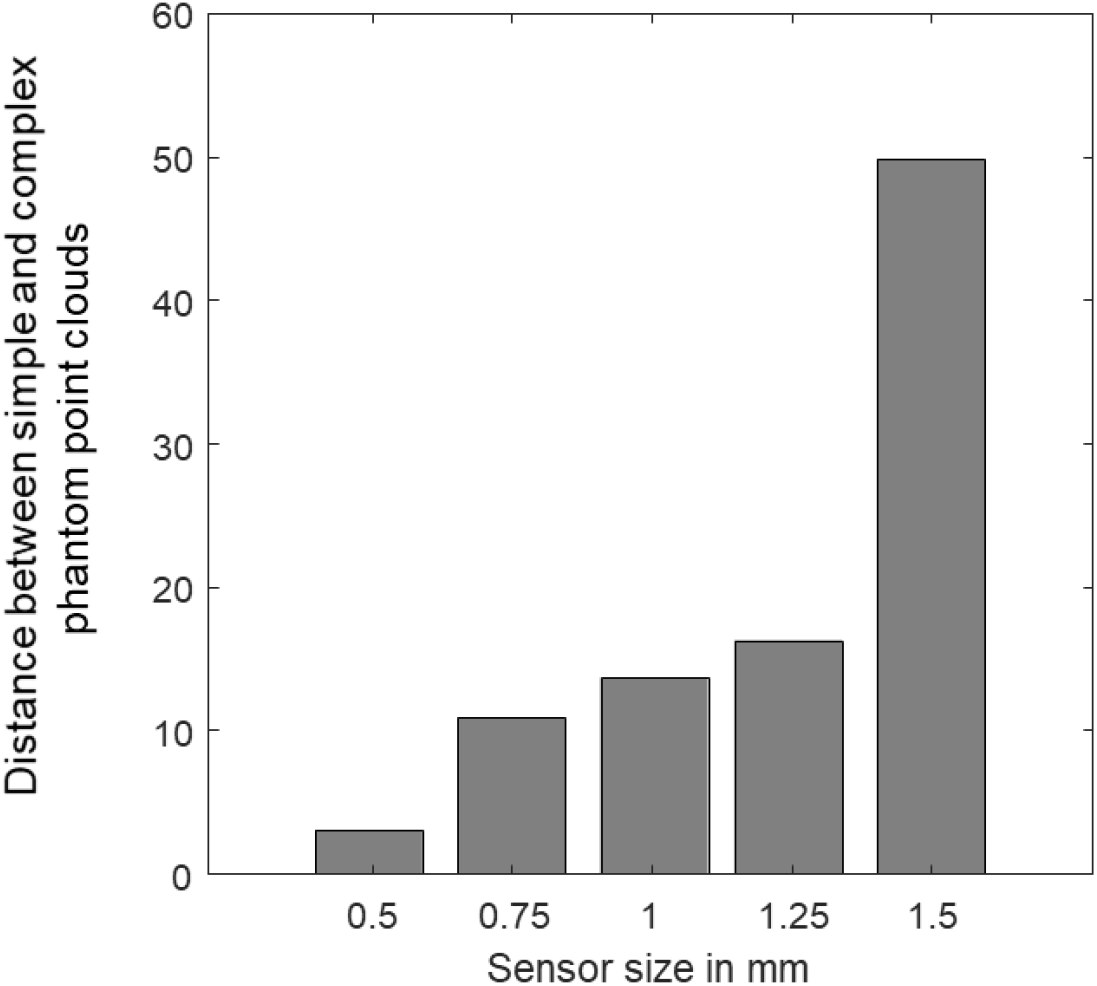
The distance between the point cloud of the simple phantom and the point cloud of the complex phantom in the t-SNE space (respectively, gray and red dots in Fig. 2) vs sensor size.

Supp. Fig. 2 presents the distance between the t-SNE points clouds of the simple and complex phantoms. For two groups of points The separation between point clouds increases with sensor size; suggesting better differentiability for larger sensors. This observation is further confirmed for clinical data.

The normalized distance between 2-D point sets *A* and *B* is denoted by *d(A,B)* and is defined as:

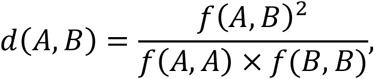

where *f(A,B)* is the average distance between a point in *A* from all the points in *B*:

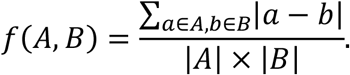

For completeness, we also provide herein the ROC curves for the trained classifiers. Note that since we have a 3-class problem, ROC curves are generated in one-vs-rest fashion: once for healthy vs rest (separately for each of the three Cases) and once for diabetes with disease vs rest. The ROC curves for the former and later cases are presented in Supp. Fig. 3, respectively. The corresponding area under curve (AUC) values are presented in Supp. Table 1.

**Supplemental Figure 3.**
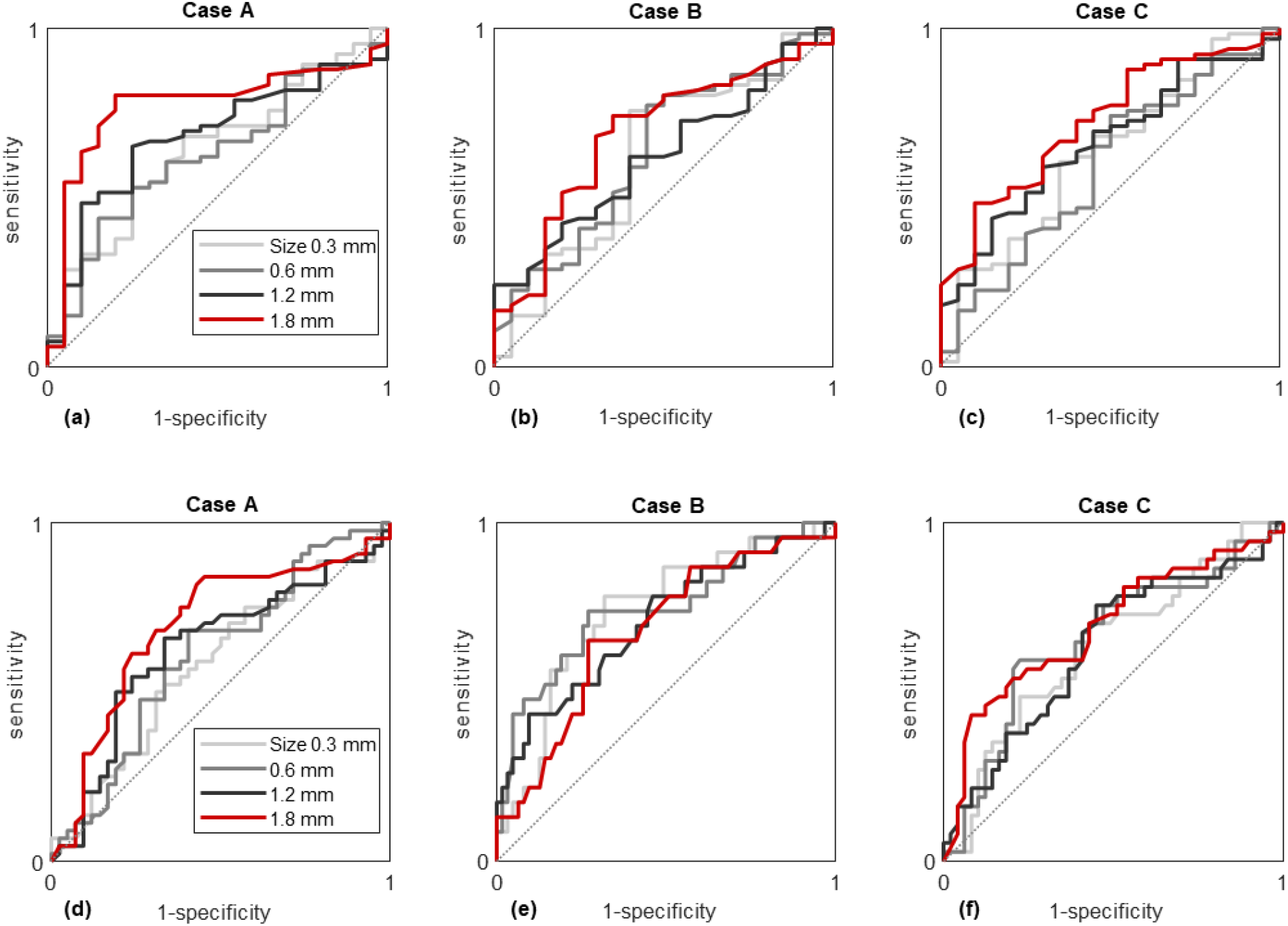
Receiver operator characteristic (ROC) curves presented for the trained classifiers as sensitivity vs 1-specificity. The three upper panels (a,b,c), correspond to, respectively, Cases A, B and C for the binary classification problem of healthy vs. rest (including diabetes with and without disease). The ROC curves are shown for different sensor sizes, as specified by the legend in panel a. Panels d, e and f show the ROC curves for the binary classification problem of diabetes with disease vs. rest (healthy and diabetes without disease).

**Supp. Table 1.**
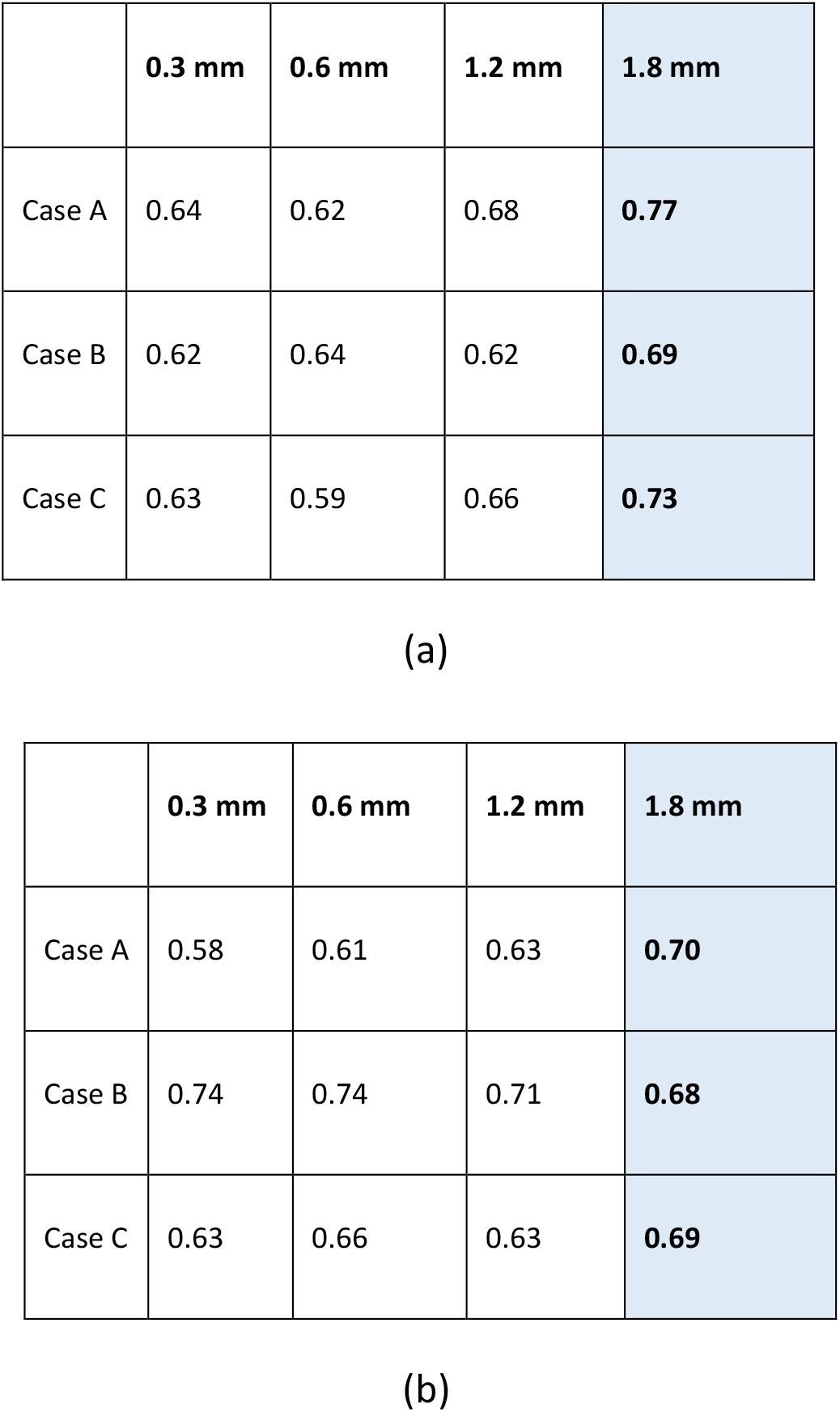
Area under curve (AUC) values obtained for the ROC curves presented in Supp. Fig. 3. Table (a) presents the results for the healthy vs rest and table (b) presents the results for ‘diabetes with diseases’ vs rest.

## References

1. Salim, A. and S. Lim, Recent advances in noninvasive flexible and wearable wireless biosensors. Biosensors and Bioelectronics, 2019: p. 111422.

2. Ray, T.R., et al., Bio-integrated wearable systems: a comprehensive review. Chemical reviews, 2019. 119(8): p. 5461–5533.

3. Mayer, M. and A.J. Baeumner, A Megatrend Challenging Analytical Chemistry: Biosensor and Chemosensor Concepts Ready for the Internet of Things. Chemical reviews, 2019. 119(13): p. 7996–8027.

4. Yu, Y., et al., Flexible Electrochemical Bioelectronics: The Rise of In Situ Bioanalysis. Advanced Materials, 2019: p. 1902083.

5. Ma, Y., et al., Flexible Hybrid Electronics for Digital Healthcare. Advanced Materials, 2019: p. 1902062.

6. Gao, Y., et al., Flexible Hybrid Sensors for Health Monitoring: Materials and Mechanisms to Render Wearability. Advanced Materials, 2019: p. 1902133.

7. Kim, J., et al., Wearable biosensors for healthcare monitoring. Nature biotechnology, 2019: p. 1.

8. Bohunicky, B. and S.A. Mousa, Biosensors: the new wave in cancer diagnosis. Nanotechnology, science and applications, 2011. 4: p. 1.

9. Imani, S., et al., A wearable chemical–electrophysiological hybrid biosensing system for real-time health and fitness monitoring. Nature communications, 2016. 7: p. 11650.

10. Ankhili, A., et al., Washable and reliable textile electrodes embedded into underwear fabric for electrocardiography (ECG) monitoring. Materials, 2018. 11(2): p. 256.

11. Administration, U.S.F.a.D. 2018; Available from: https://www.accessdata.fda.gov/cdrh_docs/pdf18/DEN180044.pdf.

12. Administration, U.S.F.a.D., 2018.

13. Heikenfeld, J., et al., Accessing analytes in biofluids for peripheral biochemical monitoring. Nature biotechnology, 2019. 37(4): p. 407–419.

14. Thennadil, S.N., et al., Comparison of glucose concentration in interstitial fluid, and capillary and venous blood during rapid changes in blood glucose levels. Diabetes Technology & Therapeutics, 2001. 3(3): p. 357–365.

15. Campbell, A.S., J. Kim, and J. Wang, Wearable electrochemical alcohol biosensors. Current opinion in electrochemistry, 2018. 10: p. 126–135.

16. Fogh-Andersen, N., et al., Composition of interstitial fluid. Clinical chemistry, 1995. 41(10): p. 1522–1525.

17. Venugopal, M., et al., Clinical evaluation of a novel interstitial fluid sensor system for remote continuous alcohol monitoring. IEEE Sensors Journal, 2008. 8(1): p. 71–80.

18. Venugopal, M., et al., A realtime and continuous assessment of cortisol in ISF using electrochemical impedance spectroscopy. Sensors and Actuators A: Physical, 2011. 172(1): p. 154–160.

19. Dias, D. and J. Paulo Silva Cunha, Wearable health devices—vital sign monitoring, systems and technologies. Sensors, 2018. 18(8): p. 2414.

20. Ntziachristos, V., et al., Looking and listening to light: the evolution of whole-body photonic imaging. Nature biotechnology, 2005. 23(3): p. 313.

21. Wang, L.V. and S. Hu, Photoacoustic tomography: in vivo imaging from organelles to organs. science, 2012. 335(6075): p. 1458–1462.

22. Pramanik, M. and L.V. Wang, Thermoacoustic and photoacoustic sensing of temperature. Journal of biomedical optics, 2009. 14(5): p. 054024.

23. Kottmann, J., et al., Mid-infrared fiber-coupled photoacoustic sensor for biomedical applications. Sensors, 2013. 13(1): p. 535–549.

24. Sörensen, B.M., et al., Prediabetes and type 2 diabetes are associated with generalized microvascular dysfunction: the Maastricht Study. Circulation, 2016. 134(18): p. 1339–1352.

25. Levy, B.I., et al., Impaired tissue perfusion: a pathology common to hypertension, obesity, and diabetes mellitus. Circulation, 2008. 118(9): p. 968–976.

26. Braverman, I.M. The cutaneous microcirculation. in Journal of Investigative Dermatology Symposium Proceedings. 2000. Elsevier.

27. Zimmet, P., K. Alberti, and J. Shaw, Global and societal implications of the diabetes epidemic. Nature, 2001. 414(6865): p. 782.

28. Das, S.K. and S.C. Elbein, The Genetic Basis of Type 2 Diabetes. Cellscience, 2006. 2(4): p. 100–131.

29. Association, A.D., 2. Classification and diagnosis of diabetes: standards of medical care in diabetes—2018. Diabetes care, 2018. 41(Supplement 1): p. S13–S27.

30. Committee, I.E., International Expert Committee report on the role of the A1C assay in the diagnosis of diabetes. Diabetes care, 2009. 32(7): p. 1327–1334.

31. Stratton, I.M., et al., Association of glycaemia with macrovascular and microvascular complications of type 2 diabetes (UKPDS 35): prospective observational study. Bmj, 2000. 321(7258): p. 405–412.

32. Omar, M., J. Gateau, and V. Ntziachristos, Raster-scan optoacoustic mesoscopy in the 25– 125 MHz range. Optics letters, 2013. 38(14): p. 2472–2474.

33. Schwarz, M., et al., Optoacoustic Dermoscopy of the Human Skin: Tuning Excitation Energy for Optimal Detection Bandwidth With Fast and Deep Imagingin vivo. IEEE transactions on medical imaging, 2017. 36(6): p. 1287–1296.

34. Miotto, R., et al., Deep learning for healthcare: review, opportunities and challenges. Briefings in bioinformatics, 2017. 19(6): p. 1236–1246.

35. Schirrmeister, R.T., et al., Deep learning with convolutional neural networks for EEG decoding and visualization. Human brain mapping, 2017. 38(11): p. 5391–5420.

36. Acharya, U.R., et al., Application of deep convolutional neural network for automated detection of myocardial infarction using ECG signals. Information Sciences, 2017. 415: p. 190–198.

37. Topol, E., Deep medicine: how artificial intelligence can make healthcare human again. 2019: Hachette UK.

38. Breiman, L., Random forests. Machine learning, 2001. 45(1): p. 5–32.

39. Bishop, C.M., Pattern Recognition and Machine Learning. 2006: Springer.

40. Maaten, L.v.d. and G. Hinton, Visualizing data using t-SNE. Journal of machine learning research, 2008. 9(Nov): p. 2579–2605.

41. Lacarrubba, F., et al., Advances in non-invasive techniques as aids to the diagnosis and monitoring of therapeutic response in plaque psoriasis: a review. International journal of dermatology, 2015. 54(6): p. 626–634.

42. Archid, R., et al., Confocal laser-scanning microscopy of capillaries in normal and psoriatic skin. Journal of biomedical optics, 2012. 17(10): p. 101511.

43. Mayrovitz, H.N. and J.A. Leedham, Laser–Doppler Imaging of Forearm Skin: Perfusion Features and Dependence of the Biological Zero on Heat-Induced Hyperemia. Microvascular research, 2001. 62(1): p. 74–78.

44. Roustit, M., et al., Excellent reproducibility of laser speckle contrast imaging to assess skin microvascular reactivity. Microvascular research, 2010. 80(3): p. 505–511.

45. Aneesh, A., et al., Multispectral in vivo three-dimensional optical coherence tomography of human skin. Journal of biomedical optics, 2010. 15(2): p. 026025.

46. Pan, Y. and D.L. Farkas, Non-invasive imaging of living human skin with dual-wavelength optical coherence tomography in two and three dimensions. Journal of biomedical optics, 1998. 3(4): p. 446–456.

47. Zabihian, B., et al., In vivo dual-modality photoacoustic and optical coherence tomography imaging of human dermatological pathologies. Biomedical optics express, 2015. 6(9): p. 3163–3178.

48. Blatter, C., et al., In situ structural and microangiographic assessment of human skin lesions with high-speed OCT. Biomedical optics express, 2012. 3(10): p. 2636–2646.

49. Errico, C., et al., Ultrafast ultrasound localization microscopy for deep super-resolution vascular imaging. Nature, 2015. 527(7579): p. 499–502.

50. Aguirre, J., et al., Precision assessment of label-free psoriasis biomarkers with ultra-broadband optoacoustic mesoscopy. Nature Biomedical Engineering, 2017. 1(5): p. 0068.

51. Omar, M., J. Aguirre, and V. Ntziachristos, Optoacoustic mesoscopy for biomedicine. Nature biomedical engineering, 2019. 3(5): p. 354–370.

52. Levy Bernard, I., et al., Impaired Tissue Perfusion. Circulation, 2008. 118(9): p. 968–976.

53. Mahe, G., et al., Assessment of skin microvascular function and dysfunction with laser speckle contrast imaging. Circ Cardiovasc Imaging, 2012. 5(1): p. 155–63.

54. Young, B.A., et al., Diabetes complications severity index and risk of mortality, hospitalization, and healthcare utilization. The American journal of managed care, 2008. 14(1): p. 15.

55. Chang, H.-Y., et al., Validating the adapted diabetes complications severity index in claims data. The American journal of managed care, 2012. 18(11): p. 721–726.

56. Ugale, S.; Available from: https://i2.wp.com/www.sages.org/wp-content/uploads/posters/60051.jpg?ssl=1.

57. Stylogiannis, A., et al., Continuous wave laser diodes enable fast optoacoustic imaging. Photoacoustics, 2018. 9: p. 31–38.

58. Nathan, D., H. Turgeon, and S. Regan, Relationship between glycated haemoglobin levels and mean glucose levels over time. Diabetologia, 2007. 50(11): p. 2239–2244.

59. Association, A.D., Introduction; Standards of Medical Care in Diabetes-2019. Diabetes Care. 42: p. S4–S6.

60. Pechenizkiy, M., A. Tsymbal, and S. Puuronen. PCA-based feature transformation for classification: issues in medical diagnostics. in Proceedings. 17th IEEE Symposium on Computer-Based Medical Systems. 2004. IEEE.

61. Latifoğlu, F., et al., Medical diagnosis of atherosclerosis from Carotid Artery Doppler Signals using principal component analysis (PCA), k-NN based weighting pre-processing and Artificial Immune Recognition System (AIRS). Journal of Biomedical Informatics, 2008. 41(1): p. 15–23.

62. Martis, R.J., U.R. Acharya, and L.C. Min, ECG beat classification using PCA, LDA, ICA and discrete wavelet transform. Biomedical Signal Processing and Control, 2013. 8(5): p. 437–448.

63. Chen, Y., et al., Deep learning-based classification of hyperspectral data. IEEE Journal of Selected topics in applied earth observations and remote sensing, 2014. 7(6): p. 2094–2107.

64. Seuret, M., et al. PCA-initialized deep neural networks applied to document image analysis. in 2017 14th IAPR international conference on document analysis and recognition (ICDAR). 2017. IEEE.

65. Mateen, M., et al., Fundus image classification using VGG-19 architecture with PCA and SVD. Symmetry, 2019. 11(1): p. 1.

66. He, H., et. al, A motion compensation algorithm for raster-scan optoacoustic mesoscopy (RSOM). In preparation.

67. Hogg, R.V. and J. Ledolter, Engineering statistics. 1987: Macmillan Pub Co.

